# Longitudinal analysis of T-cell receptor repertoires reveals persistence of antigen-driven CD4+ and CD8+ T-cell clusters in Systemic Sclerosis

**DOI:** 10.1101/2020.09.23.20190462

**Authors:** N.H. Servaas, F. Zaaraoui-Boutahar, C.G.K. Wichers, A. Ottria, E. Chouri, A.J. Affandi, S. Silva-Cardoso, M. van der Kroef, T. Carvalheiro, F. van Wijk, T.R.D.J. Radstake, A.C. Andeweg, A. Pandit

## Abstract

The T-cell receptor (TCR) is a highly polymorphic surface receptor that allows T-cells to recognize antigenic peptides presented on the major histocompatibility complex (MHC). Changes in the TCR repertoire have been observed in several autoimmune conditions, and these changes are suggested to predispose autoimmunity. Multiple lines of evidence have implied an important role for T-cells in the pathogenesis of Systemic Sclerosis (SSc), a complex autoimmune disease. One of the major questions regarding the roles of T-cells is whether expansion and activation of T-cells observed in the diseases pathogenesis is (auto)antigen driven.

To investigate the temporal TCR repertoire dynamics in SSc, we performed high-throughput sequencing of CD4+ and CD8+ TCRβ chains on longitudinal samples obtained from four SSc patients collected over a minimum of two years. Repertoire overlap analysis revealed that samples taken from the same individual over time shared a high number of TCRβ sequences, indicating a clear temporal persistence of the TCRβ repertoire in CD4+ as well as CD8+ T-cells. Moreover, the TCRβs that were found with a high frequency at one time point were also found with a high frequency at the other time points (even after almost four years), showing that frequencies of dominant TCRβs are largely consistent over time. We also show that TCRβ generation probability and observed TCR frequency are not related in SSc samples, showing that clonal expansion and persistence of TCRβs is caused by antigenic selection rather than convergent recombination. Moreover, we demonstrate that TCRβ diversity is lower in CD4+ and CD8+ T-cells from SSc patients compared to healthy memory T-cells, as SSc TCRβ repertoires are largely dominated by clonally expanded persistent TCRβ sequences. Lastly, using “Grouping of Lymphocyte Interactions by Paratope Hotspots” (GLIPH2), we identify clusters of TCRβ sequences with homologous sequences that potentially recognize the same antigens and contain TCRβs that are persist in SSc patients.

In conclusion, our results show that that CD4+ and CD8+ T-cells are highly persistent in SSc patients over time, and this persistence is likely a result from antigenic selection. Moreover, persistent TCRs form high similarity clusters with other (non-)persistent sequences, that potentially recognize the same epitopes. These data provide evidence for an (auto-)antigen driven expansion of CD4/CD8+ T-cells in SSc.

## 1. Introduction

Systemic Sclerosis (SSc) is a complex chronic autoimmune disease, characterized by vascular abnormalities and widespread fibrosis affecting the skin and internal organs[1]. Although the pathogenic mechanisms underlying SSc remain largely unknown, multiple lines of evidence imply an important role for CD4+ and CD8+ T-cells in the progression of the disease. Activated T-cells infiltrate the skin of SSc patients already in the early phase of the disease[2,3]. These infiltrating T-cells can cross-talk with fibroblasts, inducing fibroblast activation and apoptosis through secretion of pro-inflammatory cytokines and fas/fas ligand engagement[4,5]. Moreover, T-cells isolated from SSc patients undergo clonal expansion when cultured together with autologous fibroblasts, suggesting that auto-antigens presented by SSc fibroblasts can induce auto-reactive T-cell responses[6]. Apart in the from skin, peripheral blood T-cells from SSc patients also exhibit signs of activation and express activation markers, including IL-2R, HLA-DR, and CD29[7–9], and secrete pro-inflammatory and pro-fibrotic factors[10–12].

The T-cell receptor (TCR) is a highly polymorphic surface receptor that allows T-cells to recognize antigenic peptides presented on the major histocompatibility complex (MHC)[13]. CD4+ T-cells recognize peptides presented on the MHC class II complex, while CD8+ T-cells recognize peptides presented on the MHC class I complex. Classical TCRs are heterodimers consisting of a paired α- and β-chain. These chains making up the TCR are generated through somatic recombination of V (variable), D (diversity) and J (joining) gene segments accompanied by pseudorandom insertions and deletions of nucleic acids at their joining regions[14], thereby giving rise to an enormously diverse TCR repertoire in every individual. By this process of VDJ recombination, the small set of genes that encode the TCR can be used to create over 10^15^ potential TCR clonotypes[13,15]. Previous estimates of number of unique T-cells in a human range from 10^6^ to 10^11^ [16–18], meaning that every individual only carries a small fraction of the potential repertoire.

High throughput sequencing of TCR repertoires is emerging as a valuable tool to unravel the exact role of T-cells in autoimmune diseases. The TCR repertoire has been proposed to serve as diagnostic biomarker for various autoimmune diseases, and recent studies have identified disease-associated TCR sequences in autoimmune diseases including autoimmune encephalomyelitis (AE), systemic lupus erythematosus (SLE), and rheumatoid arthritis (RA)[19–21]. Moreover, changes in T-cell repertoire diversity have been suggested to predispose the pathological manifestations in RA patients[22]. Prior studies examining the TCR repertoire in SSc have shown that there is an oligoclonal expansion of T-cells in the skin, lungs and blood of SSc patients[23–25], suggesting that expanded T-cells are involved in the disease pathogenesis. However, the results from current studies examining at the TCR repertoire in SSc patients are limited as: a) they lack the use of high-throughput techniques; b) consider either CD4+ or CD8+ or unsorted T-cell populations; and c) they study T-cells obtained only from a single time point thereby providing a static snapshot of the TCR repertoire in SSc.

Two major hypotheses have been postulated to explain mechanism of the expansion of T-cells in the context of autoimmunity[26],[27]. The first hypothesis states that T-cells might expand non-specifically or by chance (bystander activation) due to chronic inflammation observed in autoimmune patients[27,28],[29]. In this case, proliferation of T-cells is induced through non-specific activation in the presence of TLR ligands and cytokines during an immune response. Due to inherent biases in the V(D)J recombination process, some TCRβ sequences are more prevalent as they have a high generation probability[30]. As a result of this bias, during bystander activation, naïve T-cell clones with TCRβs that have high generation probabilities have a larger chance of being at the site of action due to their increased prevalence, and therefore have a higher chance to expand. In this case, expansion is a result of chance. The second hypothesis states that clonal expansion in autoimmunity is driven by a chronic, specific response to (auto-)antigens that selectively skew the TCR repertoire[26]. Here expansion is driven by antigen specific selection rather than chance. It remains to be unraveled which of these two mechanisms contributes to the activation and expansion of autoreactive T-cells in SSc.

To better understand T-cell responses in SSc pathogenesis, here we investigate the temporal TCR repertoire dynamics in SSc patients. We performed high-throughput sequencing of TCRβ chains of sorted CD4+ and CD8+ non-naive T-cells isolated from longitudinal samples from four SSc patients collected over a minimum of two years.

## 2. Materials and Methods

### 2.1 Sample collection

Whole heparinized blood samples from SSc patients were obtained from the University Medical Center Utrecht. SSc patients were classified according to the ACR/EULAR criteria[31]. This study was conducted in accordance with the Declaration of Helsinki and was performed with approval of the Institutional Review Board of the University Medical Centre Utrecht, The Netherlands. The medical ethics committee of the UMC Utrecht approved the study (METC nr 13-697). All participants enrolled in the study signed informed, and patient samples were anonymized upon collection. Peripheral blood mononuclear cells (PBMCs) were isolated by density gradient centrifugation on Ficoll-PlaqueTM Plus (GE Healthcare, Uppsala, Sweden). pDCs, mDCs, B-cells and monocytes were first depleted by magnetic bead sorting using the autoMACs Pro Separator (Miltenyi Biotec, Bergisch Gladbach, Germany) according to the manufacturer’s protocol. The remaining peripheral blood lymphocytes (PBLs) were resuspended in freezing medium (80% FCS, (Sigma-Aldrich, Saint Louis, Missouri, USA) / 20% DMSO, (Sigma-Aldrich)) and stored in sterile cryovials in liquid nitrogen (−196°C) until further use. From all patients PBLs were collected at baseline (T0), at least one year after inclusion (T1, ranging from 12-19 months) and at least two years after inclusion (T2, ranging from 24-46 months) (Figure 1a).

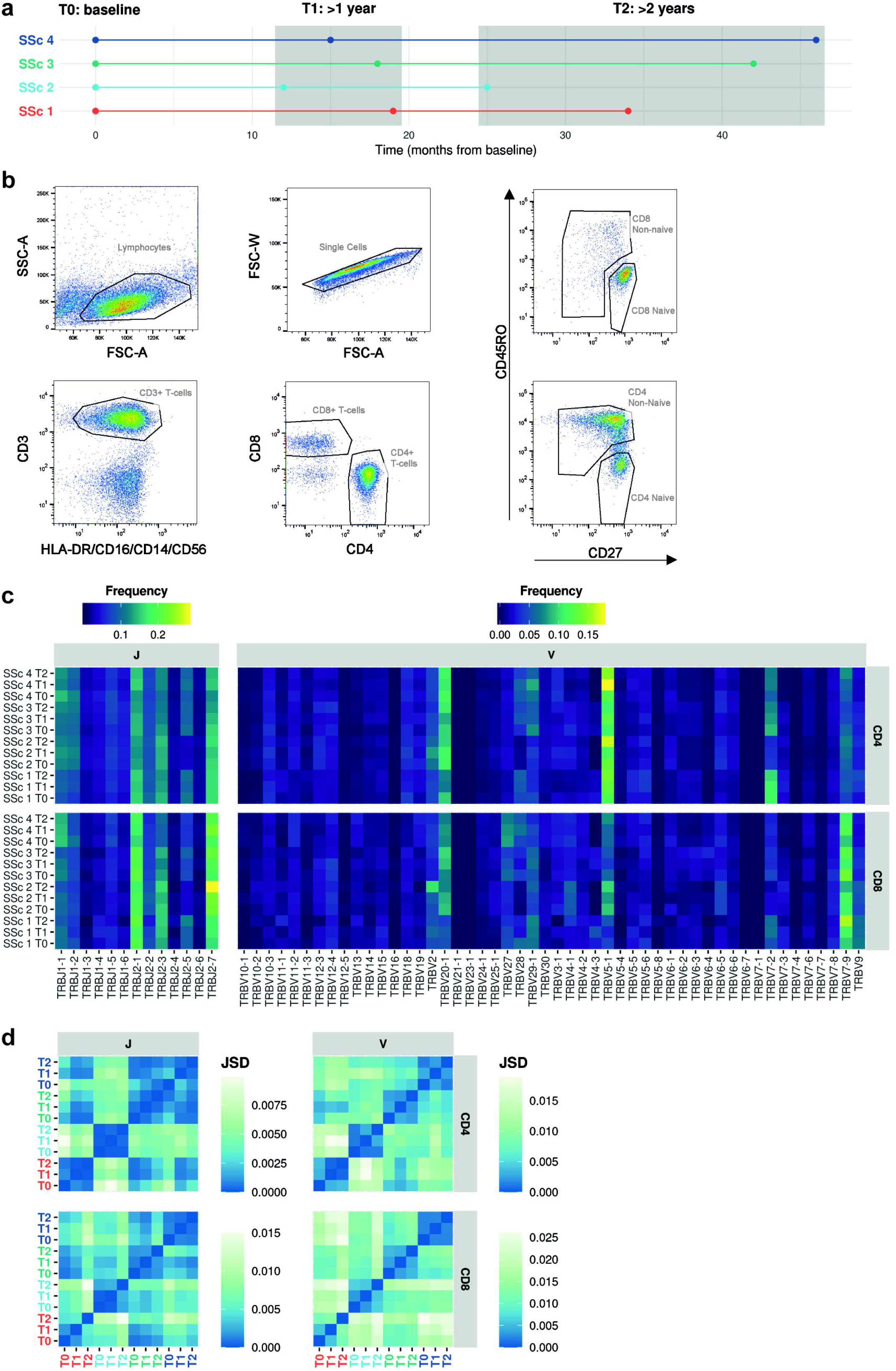

### 2.2 T-cell sorting

PBLs were thawed in RPMI 1640 (Gibco, Thermo Fisher Scientific, Waltham, Massachusetts, USA) supplemented with 20% FCS (Sigma-Aldrich), and washed with PBS. Subsequently, the cells were resuspended in FACS buffer (PBS supplemented with 1% BSA (Sigma-Aldrich) and 0.1% sodium azide (Sigma-Aldrich)) and stained using the following antibodies: CD3-AF700 (clone UCHT1, Biolegend, San Diego, California, USA), CD4-BV711 (clone OKT4, Sony Biotechnology, San Jose, California, USA), CD8-V500 (clone RPA-T8, BD Bioscience, San Jose, California, USA), CD56-PE-CF594 (clone B159, BD Bioscience), CD16-BV785 (clone 3G8, Sony Biotechnology), CD14-PerCP-Cy5.5 (clone HCD14, Biolegend), HLADR-BV421 (clone L243, Biolegend), CD45RO-PE-Cy7 (clone L243, BD Bioscience), CD27-APC-eFluor780 (clone O323, eBioscience, San Diego, California, USA). Multiparametric flow cytometry sorting of non-naive CD4+ T-cells (CD3^+^CD4^+^CD45RO^+/^CD27^+/-^) and non-naive CD8+ T-cells (CD3^+^CD8^+^CD45RO^+/-^CD27^+/-^) was performed on the BD FACSAria II (BD Bioscience), according to the gating strategy described in Figure 1b. After sorting, cells were washed in PBS, lysed in TRIzol™ Reagent (Invitrogen, Carlsbad, California, USA), and stored at −20°C until RNA isolation.

### 2.3 RNA isolation and TCR sequencing

RNA was isolated using the RNeasy Mini Kit (Qiagen, Venlo, Netherlands) by adding ethanol to the upper aqueous phase of processed TRIzol samples and transferring directly to the RNeasy spin columns. TCR amplification was performed according to the protocol published by Mamedov et al[32]. Primer and barcode sequences are provided in Supplementary Table 1. Briefly, cDNA was generated by RACE using a primer directed to the TCRβ constant region. Thirteen nucleotide long unique molecular identifiers (UMIs) were incorporated during cDNA synthesis. Subsequently, two-stage semi-nested PCR amplification was performed including a size selection/agarose gel purification step after the first PCR. To minimize cross-sample contamination, 5-nucleotide sample specific barcodes were introduced at two steps during the library preparation process[32]. Resulting TCR amplicons were subjected to high-throughput sequencing using the Ovation Low Complexity Sequencing System kit (NuGEN, San Carlos, California, USA) according to the manufacturer’s instructions, and the Illumina MiSeq system (Illumina, San Diego, California, USA), using indexed paired-end 300 cycle runs.

### 2.4 TCR repertoire analysis

Raw paired-end reads were assembled using Paired-End reAd mergeR (PEAR)[33]. Sample specific barcode correction was performed using the ‘umi_group_ec’ command from the Recover T Cell Receptor (RTCR) pipeline[34], allowing zero mismatches in the barcode seed sequence for UMI detection (sample specific barcodes are provided in Supplementary Table 1). This strict barcode selection resulted in about 50% loss of reads, but ensured that there was minimal cross-sample contamination. Subsequently, barcode sequence reads having the same UMI were collapsed into consensus sequences using the RTCR pipeline to accurately recover TCRβ sequences. Downstream data analysis of TCRβ repertoires was performed using the tcR R package[35]. Healthy longitudinal sequencing data[dataset][36] was obtained from the immuneACCESS portal of Adaptive Biotechnologies repository at: https://clients.adaptivebiotech.com/pub/healthy-adult-time-course-TCRB.

### 2.5 Statistical analyses

Statistical analyses were performed using R version 3.4.1[37], and figures were produced using the R package ggplot2[38]. Generation probabilities of TCRβ amino acid sequences were computed using the generative model of V(D)J recombination implemented by OLGA (Optimized Likelihood estimate of immunoGlobulin Amino-acid sequences)[39], using the default parameters. Diversity estimates were calculated by sample-size-based rarefaction and extrapolation using the R package iNEXT (iNterpolation/EXTrapolation)[40]. Clustering analysis was performed using the GLIPH2[41] webtool (http://50.255.35.37:8080/). Significant clusters were considered based on the following parameters: number of samples=3, number of CDR3>=3, vb_score<0.05, length_score<0.05. After filtering for significance, clusters were ordered based on final_score obtained from GLIPH2. Network graphs of clusters were produced using the R package igraph[42]. Unless indicated otherwise, analysis of differences was performed using Student t test. For multiple group comparisons, one-way anova was used. P-values <0.05 were considered statistically significant.

### 2.6 Availability of data

The TCRβ sequencing data presented in this study have been deposited in NCBI’s Gene Expression Omnibus (GEO) database under GEO: GSE156980. Both raw data and processed data are available.

## 3. Results

### 3.1 High-throughput TCR sequencing of SSc patients

To investigate the TCR repertoire dynamics in SSc, we performed high-throughput sequencing (HTS) of TCRβ chains on longitudinal samples obtained from four SSc patients collected over a minimum of two years. The clinical characteristics of the SSc patients are included in Table 1. Among the SSc patients included, two were limited SSc (lcSSc), and two were diffuse SSc patients (dcSSc). From all patients PBLs were collected at baseline (T0), at least one year after inclusion (T1, ranging from 12-19 months) and at least two years after inclusion (T2, ranging from 24-46 months) (Figure 1a). Frozen PBL samples were subjected to FACS sorting and sorted non-naive CD4+ T-cells (CD3^+^CD4^+^CD45RO^+/-^CD27^+/-^) and non-naive CD8+ T-cells (CD3^+^CD8^+^CD45RO^+/-^CD27^+/-^) were used for TCRβ sequencing (Figure 1b). Sample specific barcodes and UMIs to barcode individual mRNA molecules were used to accurately recover TCRβ sequences using the RTCR pipeline[34]. We produced a total of 906 448 and 125 962 TCRβ UMI corrected amino acid (AA) sequence reads for CD4+ and CD8+ T-cells respectively. The average number of UMI corrected AA sequence reads per sample was +/-75 000 for CD4+ T-cells and +/- 10 500 for CD8+ T-cells (details see Supplementary Table 2).

**Table 1.**
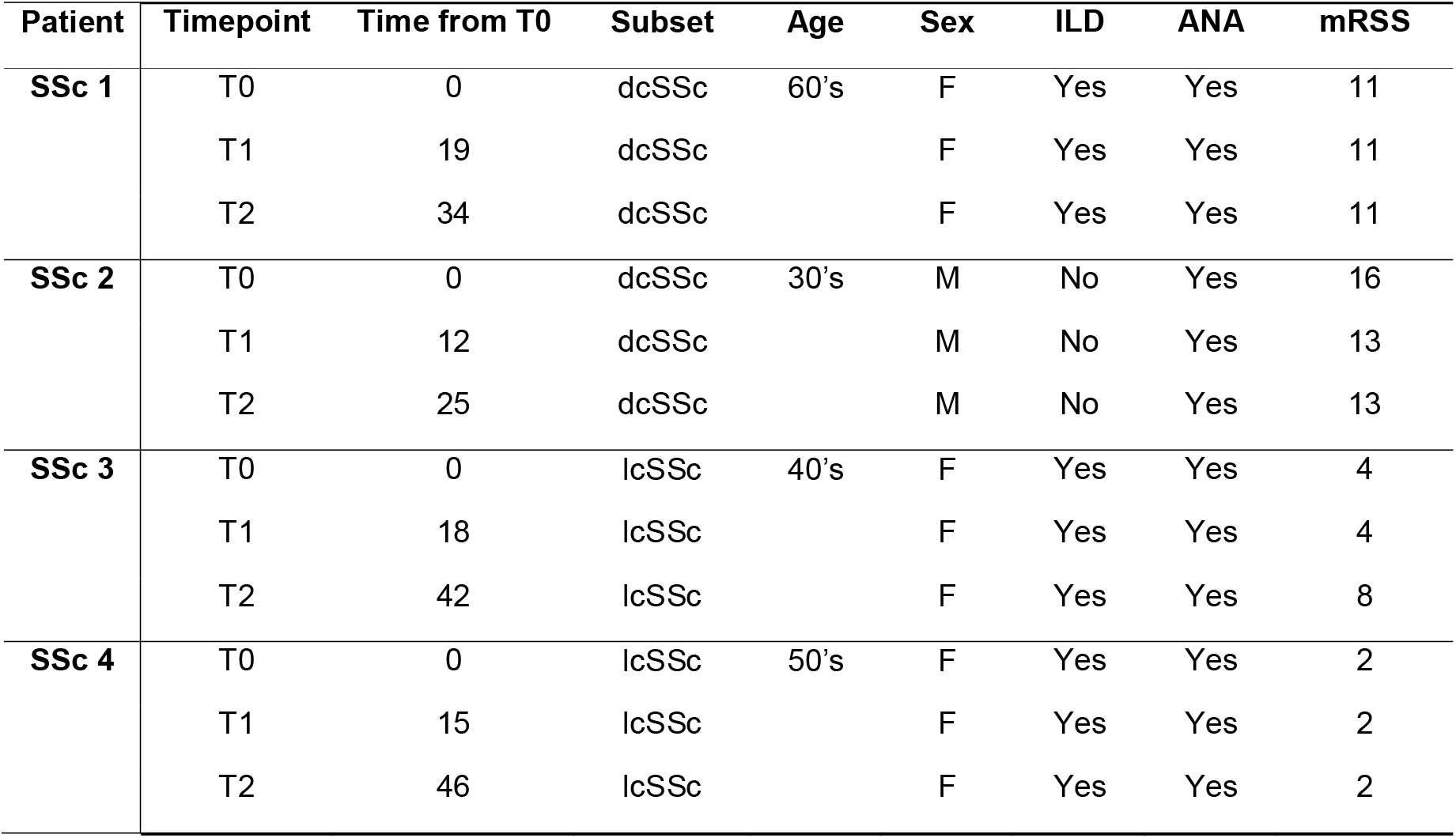
Clinical characteristics of patients included. Time from T0 is indicated in months after the first sample was taken. Abbreviations: dcSSc=diffuse SSc, lcSSc=limited SSc, M=male, F=female, ILD=interstitial lung disease, ANA= anti-nuclear antibodies, mRSS= modified rodman skin score. Age is age range between inclusion (T0) and T2.

| Patient | Timepoint | Time from T0 | Subset | Age | Sex | ILD | ANA | mRSS |
| --- | --- | --- | --- | --- | --- | --- | --- | --- |
| **SSc 1** | T0 | 0 | dcSSc | 60's | F | Yes | Yes | 11 |
|  | T1 | 19 | dcSSc |  | F | Yes | Yes | 11 |
|  | T2 | 34 | dcSSc |  | F | Yes | Yes | 11 |
| **SSc 2** | T0 | 0 | dcSSc | 30's | M | No | Yes | 16 |
|  | T1 | 12 | dcSSc |  | M | No | Yes | 13 |
|  | T2 | 25 | dcSSc |  | M | No | Yes | 13 |
| **SSc 3** | T0 | 0 | lcSSc | 40's | F | Yes | Yes | 4 |
|  | T1 | 18 | lcSSc |  | F | Yes | Yes | 4 |
|  | T2 | 42 | lcSSc |  | F | Yes | Yes | 8 |
| **SSc 4** | T0 | 0 | lcSSc | 50's | F | Yes | Yes | 2 |
|  | T1 | 15 | lcSSc |  | F | Yes | Yes | 2 |
|  | T2 | 46 | lcSSc |  | F | Yes | Yes | 2 |

### 3.2 Frequency of Vβ and Jβ gene segment usage indicates temporal persistence of TCRβ sequences in SSc patients

We first assessed the frequency of Vβ and Jβ gene segment usage in SSc patients over time (Figure 1c). The most frequently used Vβ segments across all samples were V20-1, V5-1 and V7-9, for both CD4+ and CD8+ T-cells. When looking at Jβ segment usage, J2-7, J2-1 and J2-3 were most frequently observed. In previous studies, these Vβ (V20-1, V5-1 and V7-9) and Jβ genes (J2-7, J2-1 and J2-3) were also identified as the most frequently used genes in both healthy and diseased individuals[43,44], reflecting known intrinsic biases in the V-D-J rearrangement process[45]. Additionally, Vβ2, which we identified with a relatively high frequency in CD4+ and CD8+ T-cells, was previously found to be one of the most frequent Vβ chains in peripheral blood T-cells in SSc patients in another study[46], showing that disease specific patterns are also present. Furthermore, we also observed individual specific patterns of Vβ and Jβ segments. As an example, SSc patient 2 displayed a lower frequency of V-28, V7-2 and J2-5 in both CD4+ and CD8+ T-cells across all time points compared to the other patients (Figure 1c).

In order to quantify the relative similarity of Vβ and Jβ gene segment usage across all samples, we calculated the Jensen-Shannon divergence (JSD) between them (Figure 1d). The JSD ranges from 0-1. A JSD of 0 indicates identical Vβ and Jβ segment usage, while a JSD of 1 indicates that the Vβ and Jβ segment usage is distinct between two samples. When comparing the JSD between samples taken from the same patient, we observed that the use of Vβ and Jβ segments for both CD4+ and CD8+ T-cells was rather consistent over time, while samples taken from different patients displayed a higher divergence amongst each other (Figure 1d). Moreover, when comparing the differences in JSD between Vβ and Jβ segment usage, we observed that Vβ usage (Figure 1d, right panels) was more variable then Jβ segment usage (Figure 1d, left panels) across different individuals. This difference in variability between Vβ and Jβ usage is to be expected since the TCRβ locus has more Vβ than Jβ gene segments (according to the ImMunoGeneTics (IMGT) database[47]), resulting in a greater potential variability for Vβ segment usage. Overall this analysis shows that, Vβ and Jβ gene segment usage is largely persistent within SSc patients over time.

### 3.3 SSc TCRβ repertoires are highly stable over time

Next to examining persistence in the use of Vβ and Jβ segments, we wanted to further examine the persistence of full CDR3 amino acid sequences within SSc patients over time. In order to quantify the overlap in TCRβ repertoire between different samples, Morisita’s overlap index was calculated to intersect amino acid CDR3 sequences. This index ranges from 0 (no overlapping sequences) to 1 (identical repertoires). Overlap analysis revealed that samples taken from the same patient shared a high number of sequences, indicating a clear temporal persistence of the repertoire within patients, while overlap was extremely limited between samples taken from different patients. This pattern was consistent over all time points, for both CD4+ and CD8+ T-cells (Figure 2a and Figure 2b respectively). These results demonstrate that the TCRβ repertoire in SSc patients is highly unique and stable over long periods of time, also at the level of exact CDR3 amino acid sequences.

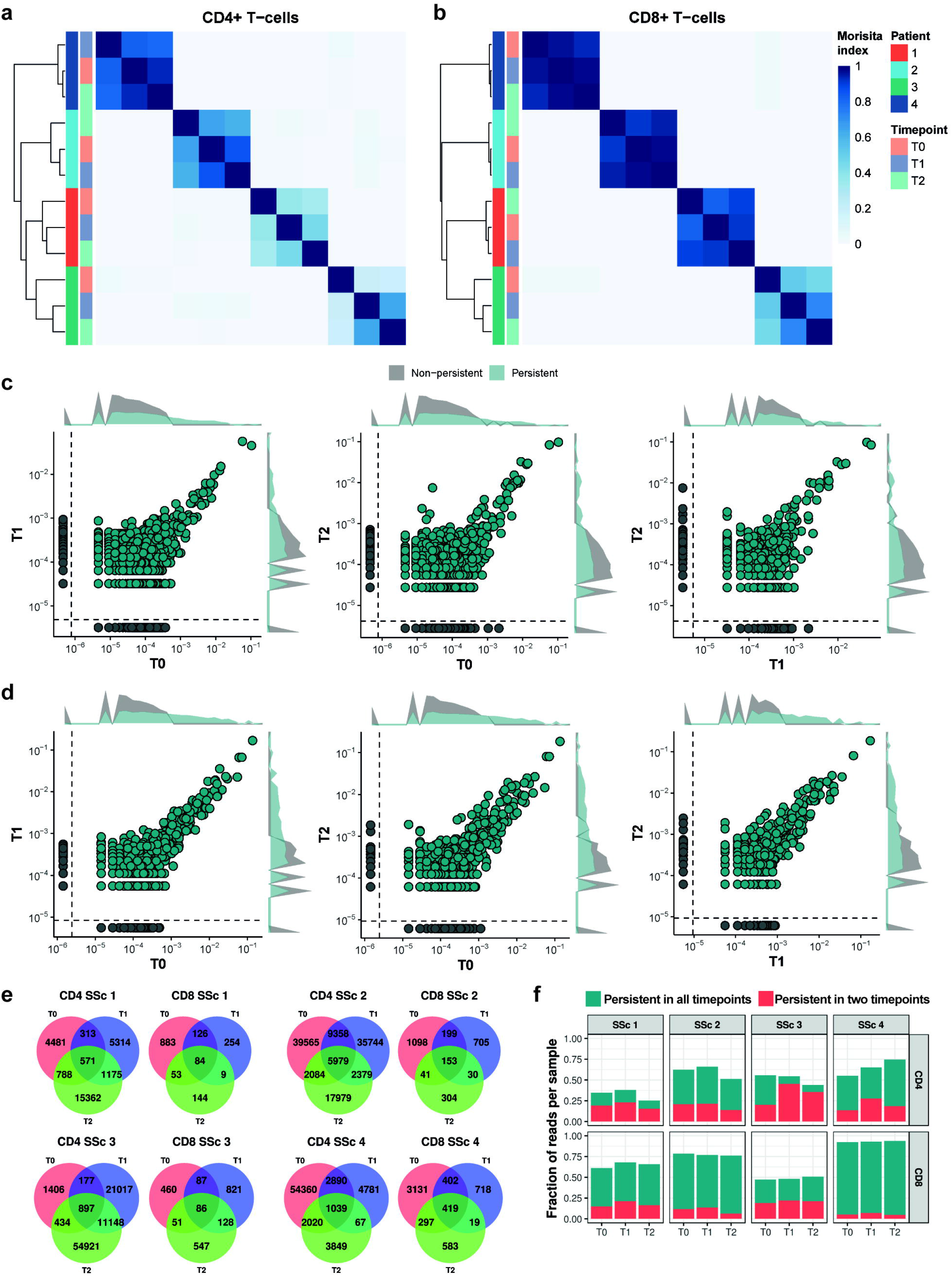

Next we examined whether the frequencies of TCRβ sequences were also consistent over time. We observed that TCRβ sequences that were found with a high frequency at one time point were also found with a high frequency at the other time points collected from the same patient, for both CD4+ and CD8+ T-cells (Figure 2c and Figure 2d respectively). This shows that the frequencies of highly abundant TCRβs within SSc patients are largely consistent over long periods of time, even after almost four years of follow-up for SSc patient 4. Persistence of dominant TCRβ sequences was observed for all the SSc patients included in our study, for both CD4+ and CD8+ T-cells (Supplementary Figure S1). The exact number of TCRβ amino acid sequences overlapping between samples taken from the same patient are shown in Figure 2e. Although the absolute number of sequences that are overlapping within patients over time (i.e. persistent sequences) are low, they make up a substantial part of the samples in terms of abundance, based on UMI corrected reads, as shown in Figure 2f. These results clearly demonstrate that SSc repertoires are largely dominated by persistent sequences.

In order to investigate whether persistent TCRβ sequences have any known antigen specificity, we queried the sequences that were persistently present in all three time points for every patient in VDJdb (a curated database of TCR sequences with known antigen specificities)[48]. The results of this analysis are shown in Table 2. In this table we show the hits for peptides presented on MHC II for CD4+ T-cells and peptides presented on MHC I for CD8+ T-cells. Hits were also found for peptides presented on MHC I for CD4+ T-cells and peptides presented on MHC II for CD8+ T-cells, even though technically these peptides cannot be recognized due to MHC restriction (Supplementary Table 3). For persistent TCRβs from CD4+ T-cells, we found most sequences to be associated with peptides presented on MHC I (Supplementary Table 3). Since CD4+ T-cells only recognize peptides presented on MHC II, these findings probably do not represent the true antigenic specificity of the TCRβ sequences queried. In general, the majority of the records present in VDJdb are based on TCR sequences obtained from MHC class I multimer assays.

**Table 2.**
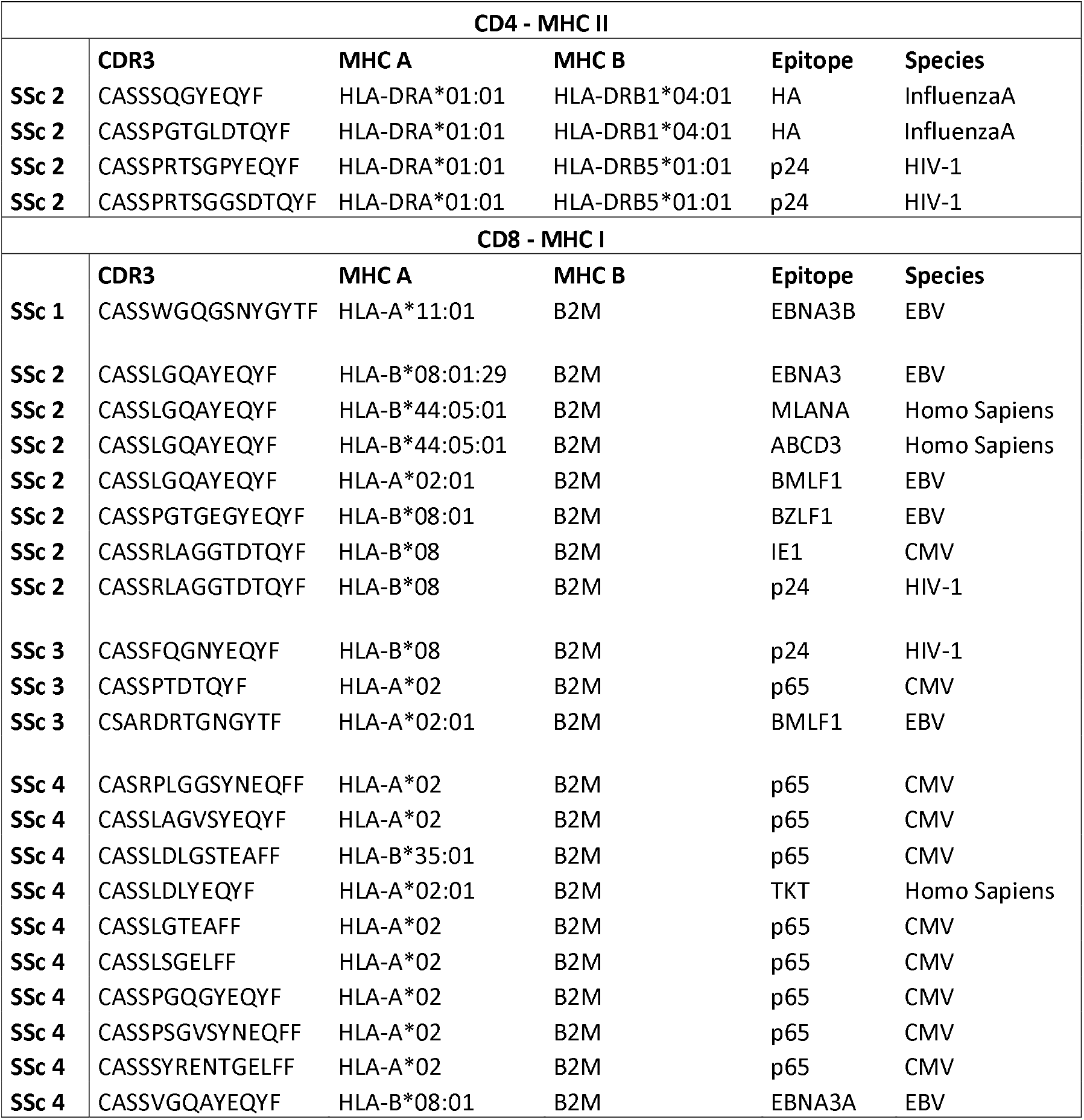
Epitope specificity of persistent TCRβs (occurring in all three time points within a patient), according to VDJdb. For CD4+ TCRβ sequences, specificities for peptides presented on MHC II are given and for CD8+ TCRβ sequences, specificities for peptides presented on MHC I are given.

For persistent TCRβs from CD8+ T-cells, we mainly found associations with peptides related to viral antigens including EBV, CMV and HIV (Table 2). For patient 2 and 3, TCRβ sequences reactive against HIV-1 epitope B2M were identified, although these patients are not HIV positive. In SSc patient 2 we also identified one TCRβ sequence (CASSLGQAYEQYF) that is associated with two human antigens, namely MLANA (melanoma antigen) and ABCD3 (ATP binding cassette subfamily D member 3). In patient 4 we identified one TCRβ sequence (CASSLDLYEQYF) that is associated with the human antigen TKT (transketolase). Interestingly, MLANA is a known antigen that is widely expressed in skin, and the presence of MLANA–specific CD8+ T-cells has been associated with autoimmune reactions in melanoma patients treated with immune checkpoint inhibitors[49].

Overall, these results reveal a clear temporal persistence of clonally expanded CD4+ and CD8+ T-cells in SSc patients. These longitudinal dynamics show that the TCRβ repertoire in SSc patients is highly stable over time, and this stability is potentially driven by a chronic response against (auto)antigens.

### 3.4 Persistent clones display common features across SSc patients

Previously, TCRs in the context of autoimmune disease have been associated with certain characteristics such as shorter length and a bias in V/J- gene segment usage[21,50,51]. To further investigate the potential involvement of persistent TCRβs identified in SSc patients in autoimmune responses, we computed the distribution of lengths of all the TCRβ amino acid sequences and compared the lengths of persistent and non-persistent TCRβs.LThe lengths were calculated based on both incidence (without weighing sequences by their abundance) and abundance (also taking into account the frequency of the sequences). A Gaussian distribution of lengths was observed for both persistent and non-persistent sequences, in CD4+ as well as CD8+ T-cells, when looking at incidence (Figure 3a), and distribution of lengths of sequences was similar between persistent and non-persistent sequences in all samples (two sample Kolmogorov-Smirnov tests >0.05 for all comparisons). When comparing the distribution of CDR3 lengths of persistent and non-persistent sequences based on abundance, we again observed no significant differences in the distributions neither for CD4+ nor CD8+ T-cells (two sample Kolmogorov-Smirnov tests >0.05 for all comparisons, Figure 3b). Although the CDR3 length distributions where not significantly different between persistent and non-persistent sequences, In SSc patient 1 and 2, more shorter length sequences were observed in the persistent TCRβs, while in SSc patient 4, longer sequences were found (Figure 3b). However, this skewness in lengths is mainly caused by the expansion of few dominant TCRβ sequences.

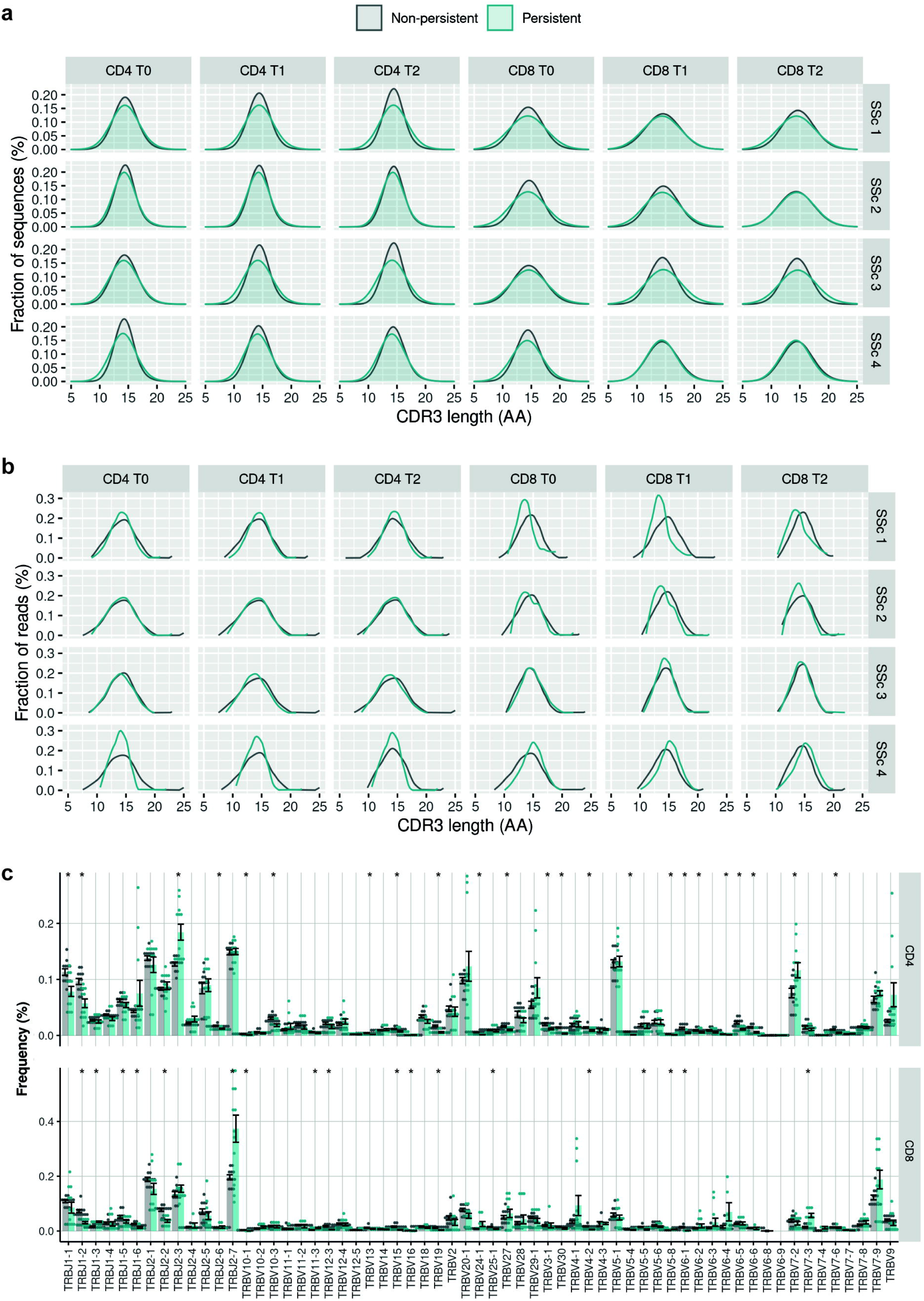

Next to comparing lengths, we also compared the frequencies of Vβ and Jβ segment usage between persistent and non-persistent TCR sequences to see if there was any preferential usage (Figure 3c). Although most differences observed were small, we identified various Vβ and Jβ gene segments that had either higher or lower frequencies in persistent sequences across all SSc patients. As an example, TRBJ1-2 had a significantly lower frequency in persistent sequences in CD4+ as well as CD8+ T-cells across all SSc patients, while the frequency of TRBV7-2 and TRBV7-3 was higher in persistent sequences in CD4+ and CD8+ T-cells respectively (Figure 3c). This analysis demonstrates that, although the number of exact sequences that are shared between SSc patients is low, TCRβ sequences that are persistently present in SSc patients over time show similarities in terms of Vβ and Jβ usage. Moreover, these are significantly different from non-persistent sequences, showing that persistent sequences display preferential usage of Vβ and Jβ segments across SSc patients. Given that similar TCR sequences are thought to be involved in T-cell responses to similar antigens[52–55], the preferential segment Vβ and Jβ segment usage across SSc patients over time might reflect chronic immune responses against antigens that are commonly present across patients.

### 3.5 SSc TCRβ repertoires are less diverse than healthy memory repertoires

The persistence of highly abundant TCRs is not necessarily unique to autoimmune repertoires and has in fact previously been observed in healthy individuals[36]. Therefore, we also compared our SSc data to a public dataset of longitudinal TCRβ sequences from healthy donors[36]. To investigate whether TCRβs in SSc patient repertoires are aberrantly expanded compared to healthy repertoires, we compared the Shannon diversities of healthy memory repertoires to those of SSc. As the samples differed in their sizes, to compare the diversity between samples we performed rarefaction analysis. The healthy control dataset from Chu *et al*. included here did not contain singletons (sequences represented by one read). We therefore performed rarefaction analysis and estimated Shannon diversity in our SSc patient data by including and excluding singletons. In Figure 4a, we show the estimated Shannon diversity based on the rarefied and extrapolated data excluding singletons. The estimated species richness and Shannon diversity was significantly lower in SSc CD4+ and CD8+ T-cell repertoires as compared to the healthy memory repertoire (p-value <0.05, Figure 4b). To estimate the effect of excluding singletons on the diversity, the same analysis was also performed including the SSc singletons. In this analysis, the estimated Shannon diversity of TCRβ sequences obtained from SSc CD8+ T-cells were still significantly lower than the diversity of healthy memory cells, while the significant difference in diversity with the CD4+ T-cells was no longer observed (Supplementary Figure S2). Overall, these results demonstrate that the TCRβ repertoire diversity is lower in SSc patients compared to healthy individuals. This provides evidence for a skewed, clonally expanded repertoire in SSc, potentially due to chronic (auto-)antigen driven T-cell responses.

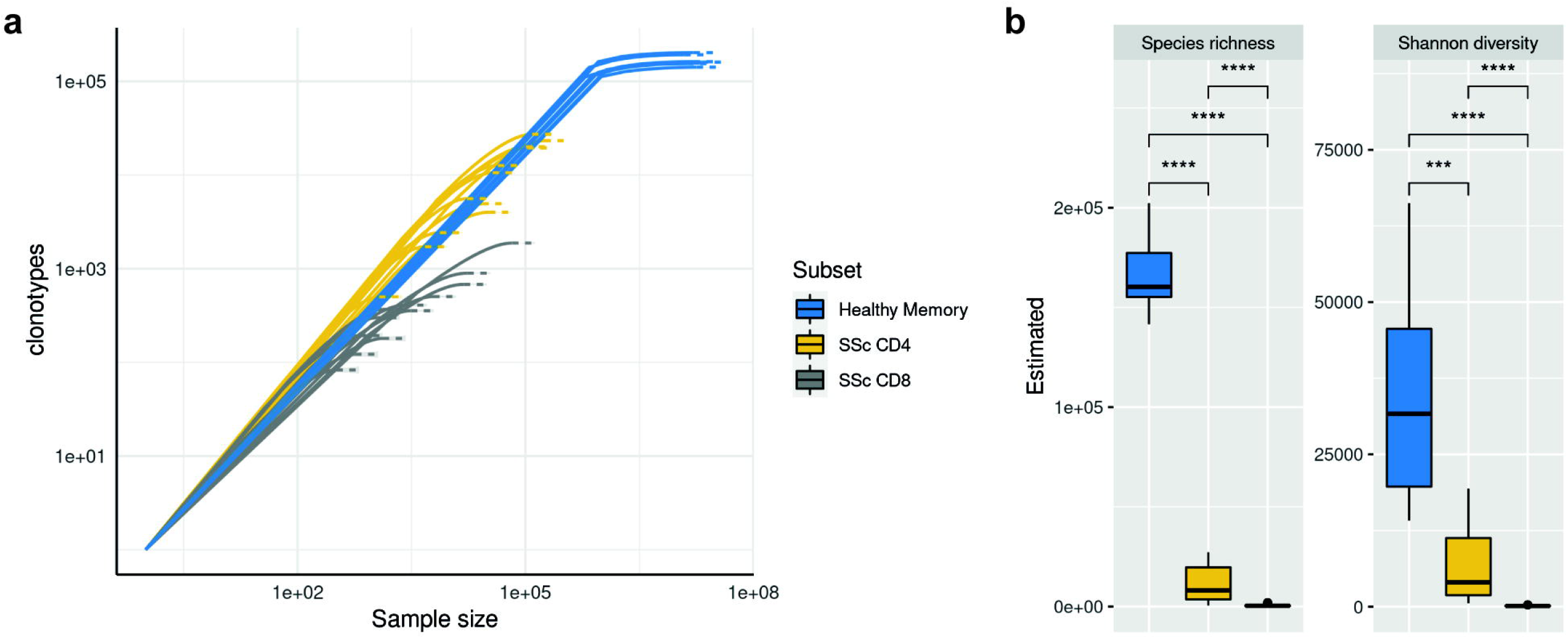

### 3.6 Persistent frequency of dominant TCR sequences is driven by antigenic selection rather than a-specific expansion

The hypothesis of 1) bystander activation due to chronic inflammation and 2) clonal expansion driven by the chronic presence of (auto-)antigens skewing the TCR repertoire have been proposed. To test the hypothesis of bystander activation, we investigated whether easy to generate TCRβ sequences (having high generation probabilities) are present at high frequencies in SSc patients. To this extend, we calculated the generation probabilities (pgens) of the TCRβs identified in SSc patients using OLGA[39]. We then used linear regression to model the relationship between TCRβ frequency and pgen. Using this analysis, we show that in SSc CD4+ and CD8+ T-cells, TCRβ frequency and pgen are not related (pvalue >0.05, Figure 5a and Figure 5b for CD4+ and CD8+ T-cells respectively). This indicates that T-cells persistent in SSc patients have not expanded because of random bystander effects.

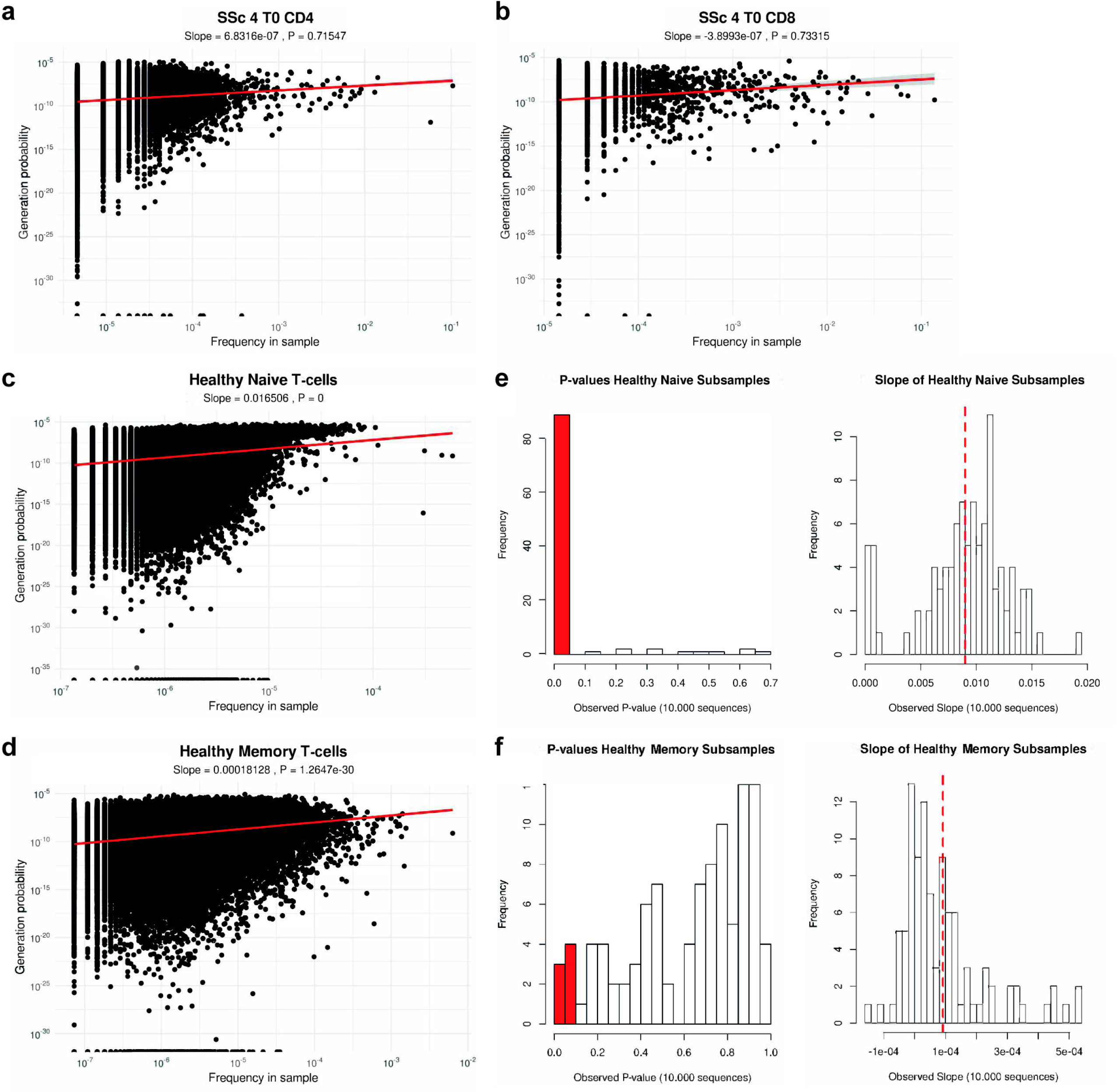

For naive T-cells isolated from healthy individuals, we found a significant positive correlation between the pgen and abundance of TCRβs (p-value <0.05, Figure 5c). For memory T-cells isolated from the same healthy individuals, we also observed a significant positive correlation between pgen and abundance of TCRβs (p-value <0.05, Figure 5d). Notably, the slope for memory T-cells was lower than that observed for the naïve T-cells (0.00018 versus 0.0165, respectively). From this analysis we show that for naïve and memory TCRβs obtained from healthy individuals, pgen and abundance are positively correlated, whereas in non-naive SSc T-cells no relationship between pgen and abundance is observed. However, as the samples obtained from healthy individuals were sequenced more deeply than our SSc samples, this observed difference in correlation might be confounded by sequencing depth. Therefore, we repeated the linear regression analysis for 100 random subsamples obtained from the healthy dataset and compared these results to the results obtained from SSc samples. Upon subsampling of the naïve healthy T-cells, 89% of the slopes observed the in linear regression were significantly different from zero (linear regression p-value <0.05 and slope >0, as indicated by the red bars in Figure 5e), confirming that indeed there is a positive relationship between pgen and abundance for TCRβs obtained from healthy naïve T-cells. However, when looking at healthy memory T-cells, we did not confirm the correlation between frequency and pgen that was observed in the full sample (p-value <0.05 in 7% of subsamples, Figure 5f), showing that in healthy memory T-cells there is no clear correlation between TCRβ pgen and abundance. Thus, subsampling analysis shows that in both healthy memory T-cells and SSc non-naïve CD4+ and CD8+ T-cells, TCRβ pgen and abundance or not correlated. These results suggest that the more abundant TCRβs in SSc repertoires are likely there because of selection, similar to what is observed in healthy memory repertoires, rather than a-specific expansion of naïve T-cells.

### 3.7 Clusters of similar TCRβ sequences can be identified in SSc patients over time

In order to identify TCRβs in SSc patients that are potentially involved in chronic autoimmune responses, we used “Grouping of Lymphocyte Interactions by Paratope Hotspots” (GLIPH2), that employs sequence similarity and motif analysis to group TCR sequences that potentially recognize the same epitope[41]. To screen for antigen specific TCR clusters, we used all sequences obtained from every time point for each individual SSc patient as an input for GLIPH2. In order to exclude false positives, for each patient we considered the clusters that contained sequences from all time points, had at least three unique CDR3s, had similar CDR3 lengths (length score <0.05), and shared similar Vβ-gene frequency distributions (Vβ score <0.05). The number of clusters that were obtained for every patient for CD4+ and CD8+ T-cells are given in Table 3.

**Table 3.**
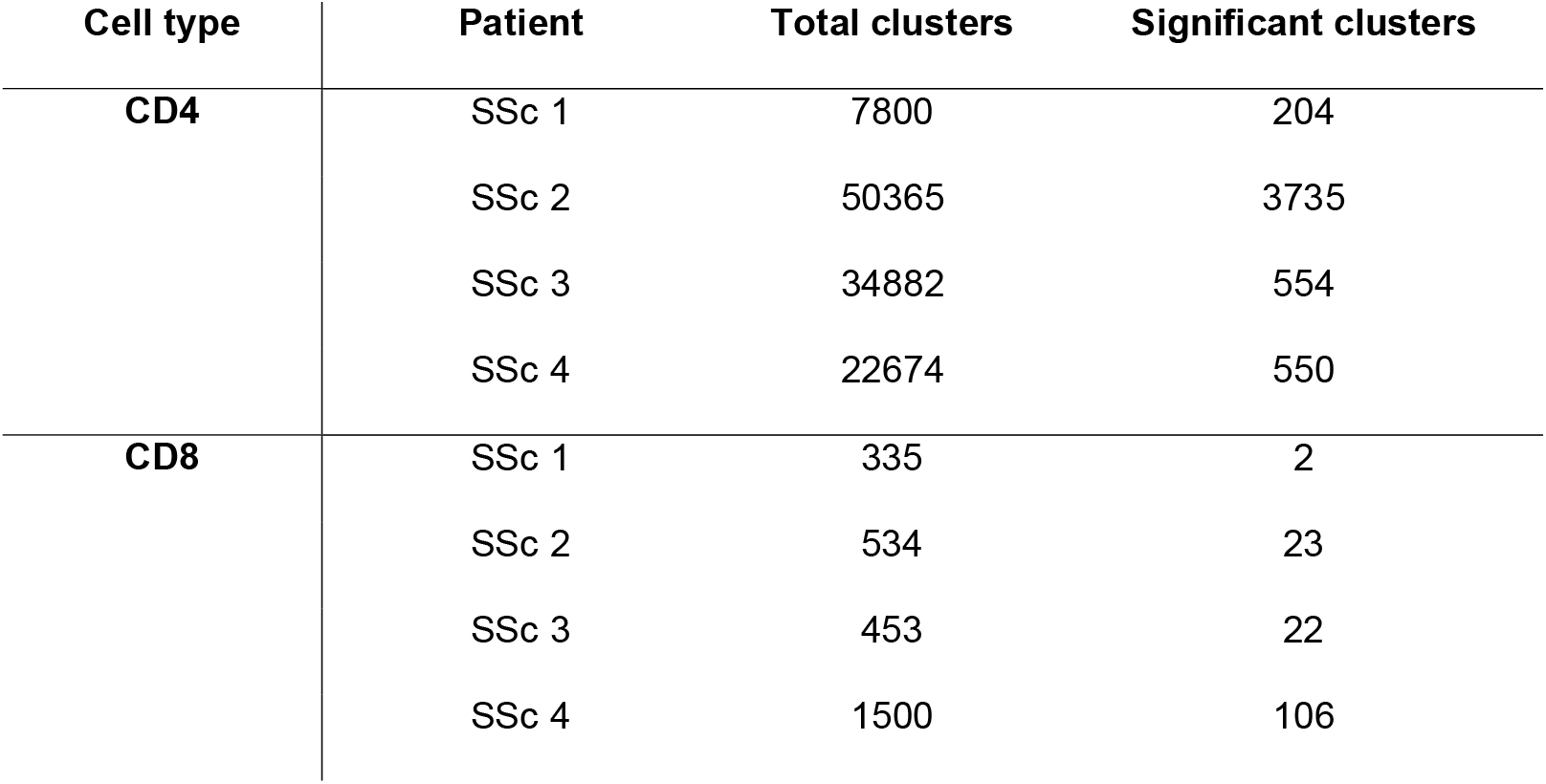
Number of clusters identified by GLIPH2 within SSc patients. All sequences from all time points from each patient (for the two cell types) were used as input for GLIPH2. The total clusters column represents the total number of clusters identified by GLIPH2. The significant clusters column represents the number of significant clusters (consisting of sequences from all three time points, at least three unique CDR3s, length score <0.05, and Vβ score <0.05).

Significant clusters were identified in all patients, either based on global similarity (CDR3 sequence differing by maximum one amino acid) or local similarity (shared motif within CDR3 amino acid region). All clusters identified by GLIPH2 are given in Supplementary Table 4. In Figure 6a, an example network of TCRβ sequences based on clustering analysis by GLIPH2 in CD4+ T-cells is given. Red and purple nodes within this network represent TCRβs that are persistently present in three or two time points respectively, while blue shaded nodes represent sequences are found only in one time point. Nodes are connected when they are part of the same cluster as identified by GLIPH2. Global and local motifs are indicated in green and orange respectively. A “%” sign within the motif indicates a variable amino acid across the sequences in which that particular motif is present.

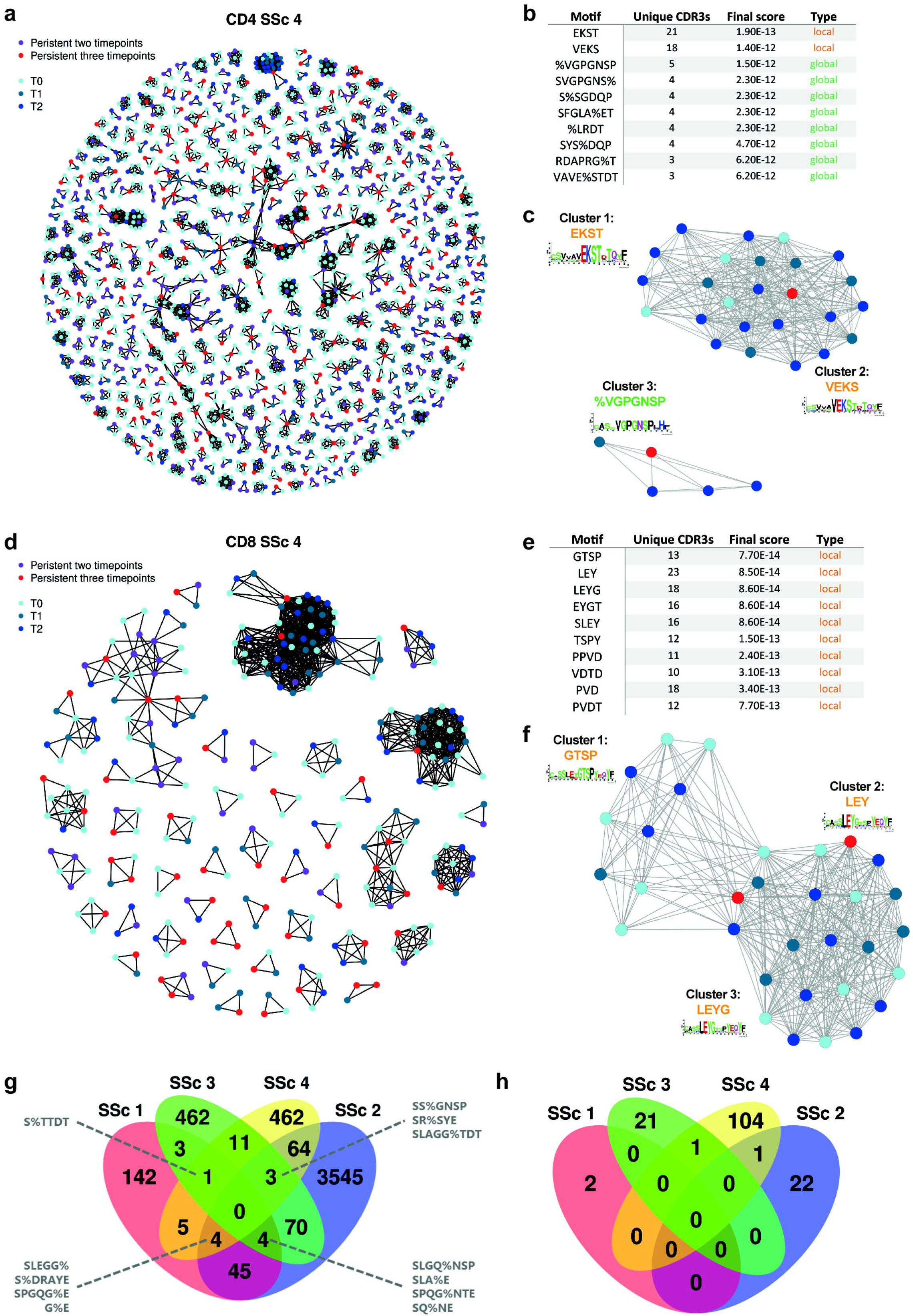

Within the network, persistent TCRβs share motifs with other persistent and non-persistent TCRβs sequences. This shows that persistent TCRβ sequences cluster together with other, similar TCRβ sequences, potentially representing groups of T-cells responding to the same antigen. The top 10 clusters, based on final cluster score outputted by GLIPH2, and their corresponding motifs for this network are given in Figure 6b. Some clusters also shared TCRβ sequences with other clusters within a patient, showing that clusters also display convergence between each other (Figure 6c, top 3 clusters are shown). Similar results were obtained for CD8+ T-cells, where we also identified many clusters of persistent and non-persistent TCRβs sharing motifs within there CDR3 region (Figure 6d-f). These results demonstrate that, apart from the presence of individual persistent clonally expanded T-cells, clusters of T-cells with potentially similar specificities are present within SSc patients over time.

Lastly, we performed an overlap analysis of all motifs from significant clusters identified by GLIPH2 between the different SSc patients (Figure 6g and Figure 6h). We did not observe any motifs for CD4+ T-cells or for CD8+ T-cells that overlap between all four patients. For the CD4+ T-cells, there were 11 motifs that were identified in clusters from three out of four SSc patients (figure 6g). These represent groups of T-cells that are likely to recognize the same or highly similar antigens across SSc patients, which could potentially be involved in SSc pathogenesis.

## 4. Discussion

Identification of TCR sequences that are associated with the chronic autoimmune response in SSc will help to get more insights into the autoimmune pathogenesis of the disease, and will help to identify the antigenic triggers that underlie these responses. Our analysis reveals that the peripheral blood TCRβ repertoire of SSc patients is highly stable over time. Moreover, the TCRβ sequences that were found within a patient with a high frequency at one time point were also found with a high frequency at the other time points (even after four years), showing that frequencies of dominant TCRβs are largely consistent over time. These persistent, clonally expanded CD4+ and CD8+ T-cells are potentially involved in the autoimmune responses underlying SSc pathogenesis. Furthermore, we have shown that the persistent expansion of these T-cells is likely a result of antigenic selection rather than recombination bias, as TCRβ frequencies were not related to their respective generation probabilities.

When we queried the persistent TCRβs found in our SSc patients in VDJdb, we obtained several hits for TCRβ sequences that are known to be associated with viral antigens from HIV, CMV and EBV, especially in the CD8+ T-cell compartment. However, as the SSc patients included in our study are all HIV negative, these hits could be potentially false positive. SSc patient 4 has a CMV and EBV positive status, and TCRβ sequences associated with CMV and EBV, as well as one TCRβ sequence (CASSLDLYEQYF) associated with the human autoantigen TKT were identified in this patient. For the other SSc patients included in this study, the CMV and EBV status are unknown. Interestingly, in SSc patient 2 we identified CASSLGQAYEQYF as a persistent sequence that is associated with EBV epitopes BMLF1 and EBNA3, as well as the human autoantigens MLANA and ABCD3. This TCRβ sequence might thus be cross-reactive to EBV and human autoantigen epitopes, potentially representing molecular mimicry. In fact, molecular mimicry between chronic viral antigens and human autoantigens has been proposed as a potential driver for autoimmune disease[56], and EBV and CMV infections have been shown to be environmental risk factors for SSc[57–60].

Chu *et al*. have previously shown that subsets of persistent TCRβs are also present within healthy individuals[36]. Thus, persistence of TCR sequences in itself is likely not just a characteristic of autoimmune related repertoires. However, the temporal dynamics of the TCR repertoire in healthy individuals in the aforementioned study have only been investigated over a period of one year, so it remains to be seen whether this stability is also maintained in healthy individuals over longer periods of time, as is observed in SSc in our study. Moreover, whereas the previous study looked into the total pool of memory T-cells, we show that persistent TCRβs can be identified in both the CD4+ and CD8+ memory T-cell compartments separately.

When further comparing the TCR repertoires of CD4+ and CD8+ T-cells from SSc patients to repertoires obtained from healthy memory T-cells, we found that SSc repertoires have lower diversity. Indeed, decreased diversities of TCRβ repertoires as compared to healthy have been observed in other autoimmune diseases[20,44,61], and have been proposed as a characteristic of autoimmune repertoires. Interestingly, in SSc, differences in the diversity of the TCR repertoire have also been observed between responders and non‐responders after autologous hematopoietic stem-cell transplantation (AHSCT, the only therapy with long‐term clinical benefit in SSc), with non‐responders having a less diverse repertoire[62]. This provides further evidence that decreased TCR repertoire diversity contributes to the autoimmune pathogenesis of SSc.

Predicting T-cell reactivity towards antigens is one of the major areas currently investigated in the field of TCR research. Since prediction of TCR binding to a specific antigen is extremely challenging, current efforts are more focused on identifying groups of TCRs that contain certain motifs within their CDR3 region[52–55]. These groups of TCRs comprise of clones that potentially respond to the same antigen. Apart from exact sharing of TCRβ sequences between samples, we also identified clusters of TCRβs that share sequence motifs and were persistent within patients. This indicates that antigen selection reshapes the TCRβ repertoire in SSc. Potential antigens could include self-antigens (such as MLANA as associated TCRβ sequences were found to be persistent within SSc patients studied here), or chronic infections with pathogens (e.g., CMV or EBV, for which we also identified associated persistent TCRβs). Interestingly, we also found clusters of TCRβs from CD4+ T-cells within patients that shared motifs with other TCRβ clusters between patients. These could represent clusters of similar TCRβs that might contribute to more public autoimmune responses underlying SSc pathogenesis. For CD8+ T-cells, we did not find any clusters overlapping between more than two patients. Notably, in general we obtained less clusters in CD8+ T-cells than we did in CD4+ T-cells. This could be due to the fact that we sequenced less CD8+ than CD4+ T-cells, and thus this difference might be explained by a difference in sequencing depth between the two cell types.

A limitation of our study is that HLA-genotype information from the SSc patients included was not obtained, so the contribution of HLA to the TCRβ clusters and shaping of the repertoire in these patients remains unknown. To validate our findings and further study the presence of potential pathogenic role of antigen specific TCRβ clusters in SSc, larger patient cohorts should be studied. In this cohort we included a limited number of patients with similar clinical characteristics which makes it difficult to account for factors such as age, sex and ethnicity influencing the immune system. Thus, studying larger longitudinal cohorts are needed to further define disease specific clusters of autoimmune associated TCRβs. Lastly, it would also be interesting to perform immune sequencing of SSc skin to see if these TCRβ clusters/motifs can be traced back in the skin (the major organ affected by the disease) of SSc patients.

## 5. Conclusion

Our data provide evidence for an (auto-)antigen driven expansion of CD4/CD8+ T-cells in SSc. We have identified clusters of T-cell clones that are highly persistent over time, and we have shown that this persistence likely is a result of antigenic selection.

## Data Availability

https://www.ncbi.nlm.nih.gov/geo/query/acc.cgi?acc=GSE156980

## Acknowledgements

We thank all the Systemic Sclerosis patients included in this study who graciously donated their time and samples. Furthermore we thank Sanne Hiddingh and the Core Flow cytometry Facility (CFF) at the UMC Utrecht for their help in sorting the T-cells samples included in this study, and Wilfred van IJcken and the Erasmus Center Biomics team (Erasmus MC Rotterdam) for Illumina sequencing. We are grateful to Anneline Hinrichs for her help with gathering of clinical data, and Rob de Boer and Peter de Greef from the Theoretical Biology & Bioinformatics department of Utrecht University for their valuable input and discussions of the data.

## Author contributions

**N.H. Servaas:** conceptualization, formal analysis, investigation, writing - original draft, writing - review & editing, visualization **F. Zaaraoui-Boutahar**: investigation, writing - review & editing **C.G.K. Wichers**: investigation, writing - review & editing **A. Ottria**: investigation, writing - review & editing **E. Chouri**: investigation, writing - review & editing **A.J. Affandi**: investigation, writing - review & editing **S. Silva-Cardoso**: investigation, writing - review & editing **M. van der Kroef**: investigation, writing - review & editing **T. Carvalheiro**: investigation, writing - review & editing **F. van Wijk**: writing - review & editing **T.R.D.J. Radstake**: writing - review & editing, funding acquisition **A.C. Andeweg**: methodology, investigation, resources, writing - review & editing **A. Pandit**: conceptualization, methodology, writing - original draft, writing - review & editing, supervision, project administration, funding acquisition. All authors critically reviewed and approved the manuscript.

